# Costs of typhoid vaccination for international travelers from the United States

**DOI:** 10.1101/2024.07.03.24309664

**Authors:** Heesoo Joo, Brian A. Maskery, Louise K. Francois Watkins, Joohyun Park, Kristina M. Angelo, Eric S. Halsey

**Author notes:** Address: U.S. Centers for Disease Control and Prevention, 1600 Clifton Road NE, MS H16-4, Atlanta, GA 30329, USA. Disclaimer: The findings and conclusions in this article are those of the authors and do not necessarily represent the official position of the U.S. Centers for Disease Control and Prevention (CDC).

## Abstract

In the United States, typhoid vaccination is recommended for international travelers to areas with a recognized risk of typhoid exposure. Using MarketScan® Commercial Database from 2016 through 2022, we estimated typhoid vaccination costs by route (injectable vs. oral) and provider setting (clinic vs. pharmacy). Of 165,930 vaccinated individuals, 99,471 received injectable and 66,459 received oral typhoid vaccines, with 88% and 17% respectively administered at clinics. Average costs for injectable vaccination were $132.91 per person [95% confidence interval (CI): $132.68–$133.13], with clinic and pharmacy costs at $136.38 [95% CI: $136.14–$136.63], and $107.45 [95% CI: $107.13–$107.77] respectively. Oral vaccination costs averaged $81.23 per person [95% CI: $81.14–$81.33], encompassing $86.61 [95% CI: $86.13– $87.10] at clinics and $80.14 [95% CI: $80.09–$80.19] at pharmacies. Out-of-pocket costs comprised 21% and 33% of total costs for injectable and oral vaccinations. These findings may inform clinical decision-making to protect international travelers’ health.

## 1. Introduction

Typhoid fever has been estimated to cause 9.2 million cases per year worldwide, resulting in 110,000 deaths in 2019, with the highest incidence rates estimated for the World Health Organization South-East Asian, Eastern Mediterranean, and African regions [1, 2]. In the United States, typhoid fever is rare, mostly linked to travel to endemic countries [3-5]. During 2017–2019, approximately 400 cases were reported annually in the United States, with over 80% associated with international travel [6]. In 2020, cases decreased to 157 due to limited international travel during the COVID-19 pandemic [3].

In the United States, the Advisory Committee on Immunization Practices recommends typhoid vaccination for international travelers to areas with a recognized risk of typhoid exposure [7]. Accordingly, the U.S. Centers for Disease Control and Prevention (CDC) recommends typhoid vaccination for those traveling to most areas in Latin America, Africa, and Asia [7-9]. Currently, two vaccines, Typhim Vi^®^ (Typhoid Vi Polysaccharide Vaccine) and Vivotif^®^ (Typhoid Vaccine Live Oral Ty21a), are available in the United States [9]. Typhim Vi^®^ can be administered to individuals at least two years old. It is a single dose (injection) and should be administered at least two weeks before travel [9]. Vivotif^®^ is suitable for individuals six years or older and requires one capsule every other day (total: four capsules) to be completed at least one week before travel [9]. Booster doses are recommended every five years for oral vaccines or every two years for injectable vaccines to maintain protection [9, 10].

While typhoid vaccination is considered a cost-effective or cost-saving strategy in some endemic areas with high incidence rates of typhoid fever [11, 12], its cost-effectiveness for U.S. residents who travel internationally remains understudied. This analysis provides the first steps toward evaluating cost-effectiveness by using claims data to estimate typhoid vaccination cost per individual by vaccination route of administration and provider setting.

## 2. Materials and methods

We identified typhoid vaccine recipients between January 1, 2016, and December 31, 2022, in the Merative® MarketScan® Commercial Database using an online tool, MarketScan® Treatment Pathways. Individuals receiving injectable typhoid vaccines were identified using the Current Procedural Terminology (CPT) code 90691 or drug product name “Typhim Vi,” while oral vaccine recipients were identified using CPT code 90690 or drug product name “Vivotif.”

We excluded individuals with (1) capitated health plans, (2) Medicare, (3) duplicate vaccine procedure code and drug name on the same day (i.e., those with MarketScan® records of receiving typhoid vaccine both at a clinic and a pharmacy) to avoid double-counting services, (4) age below vaccine-specific minimum age requirements (i.e., younger than two years for injectable and younger than six years for oral vaccine), (5) zero total typhoid vaccine payments, and (6) outlier typhoid vaccine payments using 1.5 interquartile ranges (IQR) (Appendix Figures 1 and 2). We excluded individuals enrolled in capitated health plans or Medicare due to limited payment information associated with each claim.

We categorized individuals by route of administration (injectable vs. oral) and provider setting (clinics vs. pharmacies). For those vaccinated at pharmacies, payments included only vaccine costs because pharmacies do not charge separately for administration. However, clinic payments may or may not include vaccine administration payments in addition to vaccine payments.

Thus, we divided the clinic sample into two subgroups. Sub-sample A included individuals receiving typhoid vaccine at clinics without corresponding CPT codes for vaccine administration (90460, 90461, 90471 or 90472 for injectable; 90460, 90461, 90473 or 90474 for oral) on the same day as their typhoid vaccine CPT codes or individuals who had vaccine administration codes but zero or missing total payments for vaccine administration (i.e., invalid payments). Around 11% of injectable typhoid vaccine recipients and 73% of oral typhoid vaccine recipients at clinics either lacked corresponding CPT codes for vaccine administration or had invalid vaccine administration payments (Appendix Figures 1-A and 2-A). Clinics may not charge for oral typhoid vaccine administration because patients will complete the oral vaccine schedule at home. Sub-sample B included typhoid vaccine recipients at clinics with non-zero vaccine administration payments on the same day. Additionally, for clinic sub-sample B, we excluded outliers for vaccine administration payments using 1.5 IQR. This sample division helped to distinguish between clinics that charged for expected vaccine administration and clinics that did not charge for administration.

We examined demographics and estimated the average typhoid vaccination costs per person per administration by vaccination route and provider setting, using Stata SE 16.1. P-values below 0.01 indicated rejection of the null hypothesis (i.e., no difference between groups). Categorical variables (i.e., sex, age group, and living area) were assessed using Pearson’s chi-square tests, while continuous variables (i.e., age and payments) were analyzed using two-sample t-tests with unequal variances.

Reported payments, including insurance reimbursements and patients’ out-of-pocket payments, represent vaccination costs from the payer perspective throughout the manuscript. Typhoid vaccination costs, except for those in clinic sub-sample B, included only typhoid vaccine costs. Typhoid vaccination costs for clinic sub-sample B included both typhoid vaccine and vaccine administration costs. All costs were adjusted to 2022 U.S. dollars using the Consumer Price Index for medical care [13]. A CDC reviewer determined that the project, which used only deidentified data, did not meet the regulatory definition of human subject research and did not need institutional board review.

## 3. Results

We identified 165,930 individuals who received a typhoid vaccine between 2016 and 2022 in the United States, with 99,471 (60%) receiving the injectable vaccine and 66,459 (40%) receiving the oral vaccine (Table 1). Among injectable vaccine recipients, 88% (N=87,524) received it at a clinic, while the remaining 12% (N=11,947) received it at a pharmacy. Regarding the oral vaccine, 17% (N=11,239) received it at a clinic, while 83% (N=55,220) received it at a pharmacy.

**Table 1:** Sample statistics for individuals receiving typhoid vaccine from MarketScan Commercial Claims Database using Treatment Pathways, 2016–2022.

|  | Typhim Vi® (Injectable) |  |  |  | Vivotif® (Oral) |  |  |  | P-value<sup>2,4</sup> |
| --- | --- | --- | --- | --- | --- | --- | --- | --- | --- |
|  | Clinic | Pharmacy | Total | P-value<sup>2,3</sup> | Clinic | Pharmacy | Total | P-value<sup>2,3</sup> |  |
| Sample size, No. | 87,524 | 11,947 | 99,471 | N/A | 11,239 | 55,220 | 66,459 | N/A | N/A |
| Sex, No. (%) |  |  |  |  |  |  |  |  |  |
| Female | 47,887 (54.7) | 6,503 (54.4) | 54,390 (54.7) | 0.56 | 5,996 (53.4) | 30,532 (55.3) | 36,528 (55.0) | <0.01 | 0.26 |
| Male | 39,637 (45.3) | 5,444 (45.6) | 45,081 (45.3) |  | 5,243 (46.6) | 24,688 (44.7) | 29,931 (45.0) |  |  |
| Age (years), Mean (SD) | 30.9 (17.8) | 34.7 (16.5) | 31.3 (17.7) | <0.01 | 36.3 (16.0) | 35.3 (16.4) | 35.5 (16.3) | <0.01 | <0.01 |
| Age group, No. (%) |  |  |  |  |  |  |  |  |  |
| Min. age<sup>1</sup>–18 years | 25,584 (29.2) | 2,132 (17.9) | 27,716 (27.8) | <0.01 | 1,856 (16.5) | 10,714 (19.4) | 12,570 (18.9) | <0.01 | <0.01 |
| 19–39 years | 32,079 (36.7) | 4,995 (41.8) | 37,074 (37.3) |  | 4,514 (40.2) | 21,279 (38.5) | 25,793 (38.8) |  |  |
| 40–64 years | 29,861 (34.1) | 4,820 (40.3) | 34,681 (34.9) |  | 4,869 (43.3) | 23,227 (42.1) | 28,096 (42.3) |  |  |
| Living area No. (%) |  |  |  |  |  |  |  |  |  |
| Rural | 3,505 (4.0) | 707 (5.9) | 4,212 (4.2) | <0.01 | 758 (6.8) | 2,934 (5.3) | 3,692 (5.6) | <0.01 | <0.01 |
| Non-rural | 76,489 (87.4) | 9,393 (78.6) | 85,882 (86.4) |  | 8,669 (77.1) | 45,610 (82.6) | 54,279 (81.7) |  |  |
| Missing | 7,530 (8.6) | 1,847 (15.5) | 9,377 (9.4) |  | 1,812 (16.1) | 6,676 (12.1) | 8,488 (12.8) |  |  |

Abbreviations: SD, standard deviation; N/A, not applicable
<sup>1</sup> Minimum age requirements are 2 years old for Typhim Vi® and 6 years old for Vivotif®.
<sup>2</sup> For age, p-values are calculated using the two-sample t-test with unequal variances. For all other variables, p-values are calculated using the Pearson's chi-square test.
<sup>3</sup> P-values compare sample characteristics between clinic and pharmacy samples for injectable and oral administrations, respectively.
<sup>4</sup> P-values compare sample characteristics between total injectable and total oral samples.

About 55% of typhoid vaccine recipients were female (Table 1). Injectable typhoid vaccine recipients were significantly younger (31 years) compared to oral vaccine recipients (36 years) (p<0.01). Injectable vaccine recipients at pharmacies were significantly older than recipients at clinics (35 years vs. 31 years old; p<0.01). Conversely, individuals receiving oral vaccines at pharmacies were younger than those at clinics (35 years vs. 36 years old; p<0.01).

Oral vaccine recipients were significantly more likely to have rural residency than injectable vaccine recipients (5.6% vs. 4.2%; p<0.01). Injectable vaccine recipients at pharmacies were more likely to reside in rural areas than those at clinics (5.9% vs. 4.0%; p<0.01). However, individuals receiving oral vaccines at pharmacies were less likely to reside in rural areas than those at clinics (5.3% vs. 6.8%; p<0.01).

On average, the total cost associated with injectable typhoid vaccine, including out-of-pocket and insurance costs, was $132.91 per person [95% confidence interval (CI): $132.68– $133.13] (Table 2). The average total cost to receive an injectable vaccine at a clinic was $136.38 per person [95% CI: $136.14–$136.63]. The average total costs were $116.07 [95% CI: $115.46–$116.68] for clinic sub-sample A and $138.90 [95% CI: $138.64–$139.16] for clinic sub-sample B. Costs for clinic sub-sample B included $106.25 [95% CI: $106.06–$106.45] for vaccines and $32.65 [95% CI: $32.53–$32.76] for vaccine administration. The cost per person vaccinated at pharmacies was $107.45 [95% CI: $107.13–$107.77].

**Table 2:**
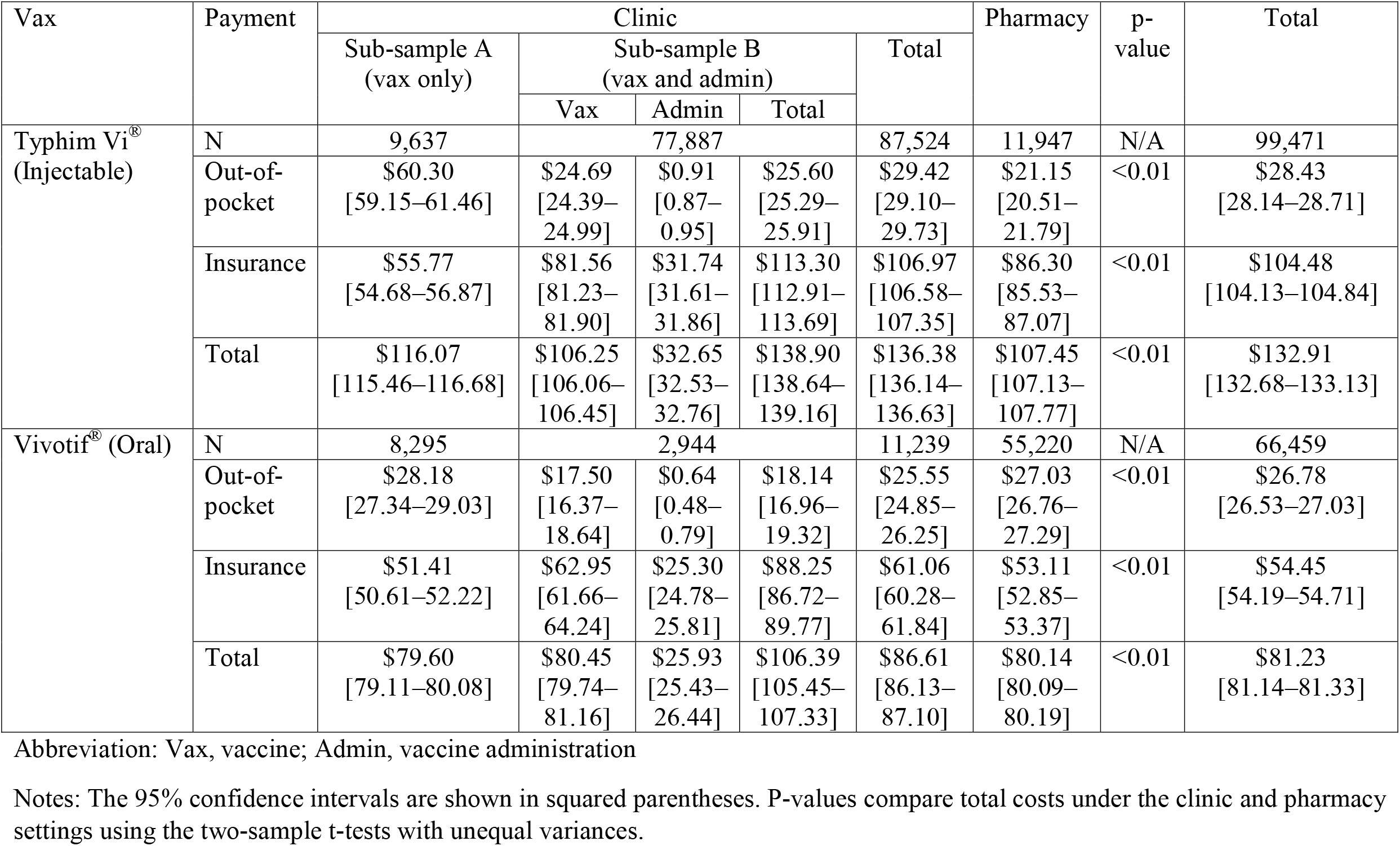
Costs associated with typhoid vaccination from the MarketScan Commercial Database in 2016–2022 (2022 USD)

The average total cost associated with oral typhoid vaccination was $81.23 per person [95% CI: $81.14–$81.33]. The cost at clinics was $86.61 per person [95% CI: $86.13–$87.10]. The cost for clinic sub-sample A was $79.60 [95% CI: $79.11–$80.08] and the cost for clinic sub-sample B was $106.39 [95% CI: $105.45–$107.33], including $80.45 [95% CI: $79.74–$81.16] for vaccine and $25.93 [95% CI: $25.43–26.44] for vaccine administration. The cost at pharmacies was $80.14 per person [95% CI: $80.09–$80.19].

The average of out-of-pocket cost associated with typhoid vaccination was $28.43 per person [95% CI: $28.14–$28.71] for an injectable vaccine and $26.78 per person [95% CI: $26.53–$27.03] for an oral vaccine (Table 2). Out-of-pocket costs constituted 21% of the injectable vaccination cost and 33% of the oral vaccination cost. The highest average out-of-pocket cost was $60.30 [95% CI: $59.15–$61.46] for clinic sub-sample A with injectable vaccine, while the lowest average out-of-pocket cost was $18.14 [95% CI: $16.96–$19.32] for clinic sub-sample B with oral vaccine.

## 4. Discussion

This study analyzed U.S. typhoid vaccination costs, which averaged around $130 per person for an injectable vaccine and $80 per person for an oral vaccine. Compared to vaccines in CDC’s adult and child/adolescent immunization schedules, which are largely covered by insurance when provided by an in-network provider [14], typhoid vaccination tends to have higher out-of-pocket costs since it is not included in these routine schedules and may not be covered by health insurance plans. This is one of the first analyses to examine U.S. costs for a vaccine that is not in the routine schedules. The high cost of typhoid vaccination and the potential that typhoid vaccines may not be covered by health insurance could pose a barrier for international travelers.

Considering vaccine cost is important during pretravel consultations. Areas with a typhoid risk may also present other infectious disease concerns, such as malaria (requiring malaria chemoprophylaxis) or yellow fever, etc. (necessitating additional vaccines). Knowing approximate out-of-pocket costs for typhoid vaccination may assist healthcare practitioners in prioritizing their recommendations.

Other factors may influence the decision to pursue typhoid vaccination. These include the waning of typhoid vaccine efficacy over time and the lack of travelers’ knowledge about travel medicine and typhoid vaccine. The estimated three-year cumulative efficacy of typhoid vaccine is around 50% for a three-dose schedule of Ty21a vaccination and 55% for a single dose of Vi polysaccharide vaccination [15]. For individuals who frequently travel to typhoid-endemic areas, typhoid vaccination may result in recurring costs due to the recommendation of booster doses, adding to their financial burden.

Vaccine availability, typhoid fever incidence rates in destination countries, the time required to complete immunization before travel, and the traveler’s medical history/age may influence the decision to recommend injectable or oral typhoid vaccine. During the COVID-19 pandemic, Vivotif^®^ was temporarily discontinued in the United States due to a substantial decrease in international travel [16]. Consequently, travelers in 2021 could have only received Typhim Vi^®^ or forego vaccination (Appendix Table 1). Travelers planning trips to areas with ongoing outbreaks or high incidence rates of typhoid fever—such as Bangladesh, India, and Pakistan [17]—may benefit more from vaccination. The time before departure may influence which vaccine is recommended to achieve optimal immune coverage during the trip [9]. If travelers visit a travel health clinic less than one week before their trip, healthcare professionals may not recommend typhoid vaccination. Also, injectable and oral typhoid vaccines have different considerations based on minimum age requirements and an individual’s medical history [9].

Individuals who received oral typhoid vaccine were older and more often lived in rural areas. Younger urban individuals may have chosen pharmacies for oral vaccine administration to avoid extra costs. Additional study is needed to clarify if vaccine cost influenced these patterns.

There are some limitations to this study. First, we did not include outpatient costs to visit a travel health clinic because travelers would typically receive guidance for several travel-associated health risks in addition to typhoid. Thus, our estimates likely underestimated the total cost. We could not determine reasons for missing vaccine administration CPT codes. Individuals lacking these codes on the day of the typhoid vaccination may have visited clinics where administration fees are not charged or may have encountered coding errors. Finally, our sample included only those with employer-sponsored private non-capitated health insurance covering typhoid vaccination, excluding entirely out-of-pocket payers, which may be common given the large number of fee-for-service U.S. travel clinics. Nevertheless, we found significant numbers of individuals who received typhoid vaccine covered by their insurance to provide an estimate of typhoid vaccination costs in the United States.

## 5. Conclusions

As international travel returns to pre-pandemic levels, the demand for travel-related vaccination, including typhoid vaccination, is anticipated to rise. Travel-related typhoid vaccination costs in the United States are crucial for informing clinical decision-making to protect international travelers’ health and evaluating cost-effectiveness of typhoid vaccination for international travelers.

## Data Availability

The data utilized for this study are accessible from a third-party source, Merative. Data availability is subject to restrictions as the data were utilized under license. Therefore, these data are not publicly accessible.

## Data availability

The data utilized for this study are accessible from a third-party source, Merative®. Data availability is subject to restrictions as the data were utilized under license. Therefore, these data are not publicly accessible.

