## Appendix for "Costs of typhoid vaccination for international travelers from the United States"

Appendix Figure 1: Sample selection for patients receiving Vi capsular polysaccharide vaccine (injectable typhoid vaccine) from MarketScan Commercial Database 2016–2022

A: Clinic Settings

Patients receiving injectable typhoid vaccine at clinic settings using procedure code 90691^1^ (n=142,127)

Patients with capitated health insurance (n=31,532)

Patients with non-capitated health insurance (n=110,595, 78%)

Patients with Medicare (n=1,460)

Patients without Medicare (n=109,135, 99%)

Duplicate exclusion: Patients with drug claims for the drug name “Typhim Vi.” on the same day as procedure code 90691 (n=219)

Patients without duplicative drug claims of “Typhim Vi.” (n=108,916, >99%)

Patients <two years old^2^ (n=547)

Patients ≥two years old
(n=108,369, >99%)

Patients with zero total payments associated with typhoid vaccine (n=13,501)

Patients with non-zero total payments associated with typhoid vaccine (n=94,868, 88%)

Patients with outlier total payments associated with typhoid vaccine ^4^ (n=4,702)

Patients without outlier total payments associated with typhoid vaccine (n=90,166, 88%)

Clinic sub-sample A
(vaccine only): Patients without vaccine administration codes or zero vaccine administration payments ^3^ (n=9,637, 11%)

Patients with vaccine
administration codes and non-zero vaccine administration payments ^3^ (n=80,529, 89%)

Patients with outlier payments associated with typhoid vaccine administration ^4^ (n=2,642, 3%)

Clinic sub-sample B
(vaccine + vaccine administration): (n=77,887, 97%)

Notes: ^1^ Patients with other injectable typhoid vaccination procedure codes (90692 or 90693) were excluded due to their extremely small sample size (N=18) and the deletion of those procedure codes effective on January 1, 2016. For individuals who receive more than one dose during the study period, we only examined the first dose received (e.g., if an individual received an initial dose in 2017 and a second dose in 2021, we dropped the observation from 2021).

^2^ We excluded patients who did not meet the minimum age requirement for injectable typhoid vaccine.

^3^ The vaccination administration fee was estimated using procedure codes 90460, 90461, 90471, or 90472.

^4^ Total payment values that were more than 1.5 times the interquartile range (IQR) above the third quartile were considered outliers.

B: Pharmacy Settings

Patients receiving injectable typhoid vaccine at pharmacy settings using the drug name “Typhim Vi.” ^1^ (n=15,191)

Patients with capitated health insurance (n=1,505)

Patients with non-capitated health insurance (n=13,686, 90%)

Patients with Medicare (n=1,188)

Patients without Medicare (n=12,498, 91%)

Duplicate exclusion: Patients with procedure code 90691 for injectable typhoid vaccination on the same day as drug claims for the drug name “Typhim Vi” (n=222)

Patients without duplicative procedure code 90691 (n=12,276, 98%)

Patients <two years old^2^ (n=13)

Patients ≥two years old
(n=12,263, >99%)

Patients with zero total payments associated with typhoid vaccine (n=86)

Patients with non-zero total payments associated with typhoid vaccine (n=12,177, 99%)

Patients with outlier payments associated with typhoid vaccine^3^ (n=230)

Study sample (n=11,947, 98%)

Notes: ^1^ For individuals who receive more than one dose during the study period, we only examined the first dose received.

^2^ Patients under two years old did not meet the minimum age requirement for injectable typhoid vaccine.

^3^ Total payment values that were more than 1.5 times the interquartile range (IQR) below the first quartile or more than 1.5 times the IQR above the third quartile were considered outliers.

Individuals who received the vaccine at a pharmacy do not have vaccination administration procedure codes.

Appendix Figure 2: Sample selection for patients receiving Ty21a vaccine (typhoid oral vaccine) from MarketScan Commercial Database 2016–2022

A: Clinic Settings^1^

Patients receiving typhoid oral vaccine at clinic settings using procedure code 90690 (n=17,735)

Patients with capitated health insurance (n=2,116)

Patients with non-capitated health insurance (n=15,619, 88%)

Patients with Medicare (n=203)

Patients without Medicare (n=15,416, 99%)

Duplicate exclusion: Patients with drug claims for the drug names “Vivotif” on the same day as procedure code 90690 (n=56)

Patients without duplicative drug claims of “Vivotif”
(n=15,360, >99%)

Patients <six years old^2^ (n=81)

Patients ≥six years old
(n=15,279, >99%)

Patients with zero total payments associated with typhoid vaccine (n=2,186)

Patients with non-zero total payments associated with typhoid vaccine (n=13,093, 86%)

Patients with outlier total payments associated with typhoid vaccine^3^ (n=1,699)

Patients without outlier total payments associated with typhoid vaccine (n=11,394, 87%)

Clinic sub-sample A (vaccine only): Patients without vaccine administration codes or zero vaccine administration payments^4^ (n=8,295, 73%)

Patients with vaccine
administration codes and non-zero vaccine administration payments^3^ (n=3,099, 27%)

Patients with outlier payments associated with typhoid vaccine administration^5^ (n=155, 5%)

Clinic sub-sample B
(vaccine +vaccine administration)
(n=2,944, 95%)

B: Pharmacy Settings

Patients receiving oral typhoid vaccine at pharmacy settings using the drug name “Vivotif”^1^ (n=79,532)

Patients with capitated health insurance (n=19,466)

Patients with non-capitated health insurance (n=60,066, 76%)

Patients with Medicare (n=2,206)

Patients without Medicare (n=57,860, 96%)

Duplicate exclusion: Patients with procedure code 90690 for oral typhoid vaccination on the same day as drug claims for the drug names “Vivotif” (n=57)

Patients without duplicative procedure code 90690
(n=57,803, >99%)

Patients <six years old^2^ (n=116)

Patients ≥six years old
(n=57,687, >99%)

Patients with zero total payments associated with typhoid vaccine (n=126)

Patients with non-zero total payments associated with typhoid vaccine (n=57,561, >99%)

Patients with outlier payments associated with typhoid vaccine^3^ (n=2,341)

Study sample (n=55,220, 96%)

Notes: ^1^ For individuals who receive more than one dose during the study period, we only examined the first dose received.

^2^ Patients under six years old did not meet the minimum age requirement for oral typhoid vaccine.

^3^ Total payment values that were more than 1.5 times the interquartile ranges (IQR) below the first quartile or more than 1.5 times the IQR above the third quartile were considered outliers.

^4^ The vaccination administration procedure codes 90460, 90461, 90473, or 90474 were used.

^5^ Total payment values that were more than 1.5 times the IQR above the third quartile were considered outliers.

Appendix Table 1: Number of individuals who received typhoid vaccine by type of vaccine, provider setting, and year

| Year | Typhim Vi^®^ (Injectable) | | | Vivotif^®^ (Oral) | | | (A)/(A+B) |
| --- | --- | --- | --- | --- | --- | --- | --- |
|  | Clinic | Pharmacy | Total, (A) | Clinic | Pharmacy | Total, (B) |  |
| 2016 | 21,389 | 2,011 | 23,400 | 3,264 | 13,270 | 16,534 | 59% |
| 2017 | 19,201 | 2,171 | 21,372 | 2,545 | 12,478 | 15,023 | 59% |
| 2018 | 18,594 | 2,589 | 21,183 | 2,519 | 14,292 | 16,811 | 56% |
| 2019 | 15,028 | 2,451 | 17,479 | 2,092 | 11,377 | 13,469 | 56% |
| 2020 | 2,613 | 421 | 3,034 | 393 | 2,345 | 2,738 | 53% |
| 2021 | 3,131 | 537 | 3,668 | 7 | 17 | 24 | 99% |
| 2022 | 7,568 | 1,767 | 9,335 | 419 | 1,441 | 1,860 | 83% |
| Total | 87,524 | 11,947 | 99,471 | 11,239 | 55,220 | 66,459 | 60% |

Notes: The number of individuals receiving oral typhoid vaccine decreased significantly in 2021 because of the temporary discontinuation of Vivotif^®^ in the United States due to the COVID-19 pandemic. The shortage due to the temporary discontinuation was resolved in June 2022.

Appendix Table 2: Costs associated with typhoid vaccination by region from the MarketScan Commercial Database in 2016–2022 (2022 USD)

|  | | Rural | Non-rural | p-value |
| --- | --- | --- | --- | --- |
| Total | Out-of-pocket | $28.47 [27.60–29.34] | $28.55 [28.32–28.77] | 0.88 |
|  | Insurance | $75.97 [74.82–77.11] | $85.45 [85.16–85.74] | <0.01 |
|  | Total | $104.44 [103.72–105.16] | $113.99[113.78–114.20] | <0.01 |
|  | N | 7,904 | 140,161 |  |
| Injectable | Out-of-pocket | $24.45 [23.17–25.73] | $29.97 [29.66–30.28] | <0.01 |
|  | Insurance | $100.87 [99.32–102.41] | $104.58 [104.19–104.97] | <0.01 |
|  | Total | $125.31 [124.40–126.22] | $134.12 [134.31–134.80] | <0.01 |
|  | N | 4,212 | 85,882 |  |
| Oral | Out-of-pocket | $33.07 [31.92–34.21] | $26.29 [26.02–26.57] | <0.01 |
|  | Insurance | $47.56 [46.40–48.72] | $55.17 [54.89–55.45] | <0.01 |
|  | Total | $80.63 [80.22–81.03] | $81.46 [81.35–81.56] | <0.01 |
|  | N | 3,692 | 54,279 |  |

Notes: The 95% confidence intervals are shown in squared parentheses. P-values are from two-sample t-tests with unequal variances.
